# Effects of a school-based physical activity implementation program to reduce musculoskeletal pain frequency in children aged 9 to 12: a randomised clinical trial

**DOI:** 10.1101/2024.01.20.24301537

**Authors:** Priscilla Viana da Silva, Steven J Kamper, Tiê Parma Yamato, Luke Wolfenden, Rachel Sutherland, Nicole McCarthy, Erin Nolan, Christopher Oldmeadow, Nicole Nathan, Christopher M Williams

## Abstract

**Objective:** To estimate the effect of scheduling physical activity during the school day on pain outcomes (frequency, severity, and impact), quality of life, physical activity and sedentary time in school children with musculoskeletal pain.

**Design, setting, and participants:** A preplanned secondary analysis of a cluster randomised trial in private elementary schools from Australia. Eligible participants were students aged 9 to 12 (grades 4 to 6) who reported having previous MSK pain.

**Interventions:** Intervention schools received support to increase the scheduling of physical activity during the school day (with and without nutrition strategies). Control schools received either support to implement nutrition strategies or no active support.

**Main outcomes and measures:** Pain frequency reported at 9-months after randomisation over the last six months. Secondary outcomes: quality of life (PedsQL 4.0), pain severity (FSP-R), physical activity and sedentary times, the prevalence of recent pain, and pain-related impact on physical activity, usual activities, school absenteeism, and health care utilisation.

**Results:** We included 633 students (mean age 10.5 years). Of those, 33% of students reported previous frequent pain at follow-up. We found no difference in pain frequency between groups (OR 1.0, 95% CI: 0.6 to 1.6). The intervention group had slightly higher physical activity levels (mean difference: 3.1 min/day, 95% CI: −0.02 to 6.2) and lower pain impact in usual activities (OR 0.7, 95% CI: 0.4 to 1.0).

**Conclusion and relevance:** Increased scheduling of physical activity in schools did not improve pain outcomes in children with MSK pain. However, results suggest that children can improve physical activity levels during school time despite experiencing pain.

**Trial registration:** ACTRN12616001228471

**SUMMARY BOX:** *What is already known on this topic:* Musculoskeletal (MSK) pain is prevalent among children and adolescents. The relationship between physical activity and pain is complex, with evidence indicating that physical inactivity can contribute to MSK pain, but pain may also serve as a barrier to engaging in physical activities.

*What this study adds:* We found that scheduling physical activity during school time did not improve pain outcomes of school-aged children with MSK pain. However, there appears to be a small benefit to improving physical activity levels and impact on usual activities in students with prior MSK pain.

*How this study might affect research, practice or policy:* This study highlights that children with MSK pain might need a more comprehensive approach to school-based physical activity programs, combining strategies that not only promote physical activity but also address the unique challenges posed by MSK pain in this age group.

## BACKGROUND

Musculoskeletal (MSK) pain is common and burdensome for children and adolescents. Between 4 and 40% of children and adolescents are affected by MSK pain globally.[1] Around 32% of children aged 9-11 experience MSK pain over a 3-month period and 40% of children aged 8-18 report MSK pain at some point during the last 6 months.[1, 2] According to the latest Global Burden of Disease study, MSK pain in people aged 10 to 24 years increased by 13.1% in the last decade, and low back pain is one of the top ten causes of MSK pain.[3] MSK pain in childhood can impact children’s daily activities.[4, 5] For example, frequent pain can lead to school absenteeism,[6] decreased social interaction and quality of life,[7, 8] and activity limitations.[9] Importantly, the impact of MSK pain may track into adulthood, suggesting that mitigating pain and its impact on childhood could have lifelong consequences.[10]

The interaction between physical activity and pain in children is uncertain. Previous studies show that physical inactivity increases the risk of low back pain, joint pain and inferior joint development.[11–13] Current evidence shows that exercise therapy is a promising strategy to improve MSK pain in children.[14, 15] However, pain may also be an important barrier for children to engage in physical activities.[16] Consequently, children with frequent MSK pain who are physically inactive may be at higher risk of obesity, diabetes and cardiovascular diseases.[11, 12] Therefore, developing strategies to support physical activity for pain management is important to ensure the health of children who experience MSK pain.[16]

Schools are recognised as an ideal setting to improve young people’s health behaviours, as they provide access to almost all children for a prolonged period of time each day.[17] Evidence shows that physical activity levels of children can be increased when they are exposed, during school hours, to quality physical education (PE), active recess and lunch, and to energisers (physical activity integrated into classroom routines).[18–20] Therefore, the Global Action of Plan on Physical Activity[21] recommends the implementation of physical activity policies in schools.

Strategies to increase physical activity time and level are not routinely implemented in schools.[22] Therefore, governments internationally have recommended or mandated schools to implement a minimum time and intensity of daily physical activity to comply with the Global Action of Plan on Physical Activity.[23–25] In New South Wales (NSW) Australia, the government mandates 150 minutes of weekly physical activity during school hours.[26] Our recent cluster randomised trial aimed to identify the most effective strategies to support NSW schools to implement this policy.[27] At follow-up we found that intervention schools teachers scheduled on average 36.6 (95% CI, 2.68–70.51) more minutes of physical activity across the school week compared to control school teachers. This result means an approximate addition of 7 more minutes of physical activity per day during school hours. Device-based data collected from more than 1500 children in kindergarten to grade 6 (aged 5 to 12) translated this finding to an increase of over 3 minutes per school day (3.0; 95% CI: 2.2 to 3.8) in moderate-to-vigorous physical activity (MVPA) with larger effects in older children in grade 5 (5.1 minutes 95% CI: 3.0 to 7.2) and grade 6 (6.0 minutes; 95% CI: 3.4 to 8.6).[27]

School-based programs may support the general health of children who experience pain.[28] Given that MSK pain can affect physical activity levels, school-based programs aiming to increase physical activity levels during school time may improve MSK pain outcomes. However, there is limited evidence as to whether school-based physical activity programs improve pain or physical activity in children with MSK pain. We conducted a pre-planned[29] secondary analysis of a cluster RCT to estimate the effects of supporting schools to increase scheduling of school-time physical activity on pain frequency in children aged 9 to 12 with MSK pain. Secondary aims were to assess the effect of the program on the quality of life, pain severity, prevalence of recent pain, physical activity and sedentary time, and pain-related impact on school absenteeism, health care use and activity participation of children with MSK pain.

## METHODS

### Context

In New South Wales, Australia, Department of Education policy requires schools to schedule at least 150 minutes of MVPA across the school week for students.[26, 30] The physical activity arm of the main cluster randomised trial tested strategies to support primary school teachers to increase the scheduling of physical activity across the school day.[27] The factorial design included three intervention groups and one waitlist (no intervention) group. The physical activity[27] and nutritional[31] outcomes were published elsewhere. This study report follows the Consolidated Standards of Reporting Trials (CONSORT).

### Design, setting and eligibility

This is a secondary analysis of a cluster randomised implementation trial, conducted in twelve Catholic schools from the Hunter Region of NSW – Australia. The cluster randomised trial was approved by the Hunter New England Human Research Ethics Committee (Ref. No. 06/07/26/4.04), The University of Newcastle Ethics Committee (Ref. No. H-2008-0343), and the Maitland-Newcastle Catholic Schools Office and prospectively registered with Australian New Zealand Clinical Trials Register ACTRN12616001228471. We conducted our analysis according to a registered statistical analysis plan.[29] All Catholic primary schools that enrolled children aged 5-12 years old, had greater than 120 students, used the school mobile communication app (Skoolbag), and were not participating in other nutrition or physical activity-based research studies were eligible to participate in the study. Schools catering only for children with special needs (such as intellectual disabilities) were excluded. All schools were provided with a study information package asking for written informed consent for the school to participate. The first twelve consenting and eligible schools were enrolled in the trial. After baseline data collection, the twelve schools were allocated to one of four groups (1) physical activity support only, (2) nutrition support only, (3) both physical activity and nutrition support, and (4) waitlist. An independent statistician randomly allocated using a computerised random number generator. School staff were aware of group allocation, and data collectors were aware of allocation at follow-up but not at baseline.

### Participants

We included students that met the following criteria: parental consent to participate in data collection; were in grades 4 to 6 (aged 9 to 12); had completed a paper survey and reported they previously experienced bodily pain “often”, “once in a while” or “once or twice”. We excluded students who reported they had “never” experienced bodily pain.

### Interventions

Intervention schools were allocated to receive physical activity implementation support strategy only or the combined physical activity and nutrition implementation support strategies together (groups 1 and 3). The implementation strategies to support teachers’ scheduling of physical activity included identifying and preparing in-school champions, development of a formal implementation blueprint, educational outreach visits for staff, and distribution of educational materials.[27] The nutrition intervention consisted of nutrition guidelines and resources to the school, teacher, and students about packing a healthy lunchbox, and communication to parents/carers addressing the barriers to packing a healthy lunchbox via the Skoolbag app.[31] The intervention period lasted nine months (January to September 2017). **Figure 1** provides further information about the physical activity and nutrition interventions. The control group for this secondary analysis were schools allocated to the nutrition implementation support strategy only or waitlist control (groups 2 and 4). The waitlist group received no active support during the trial period.

**Figure 1.**
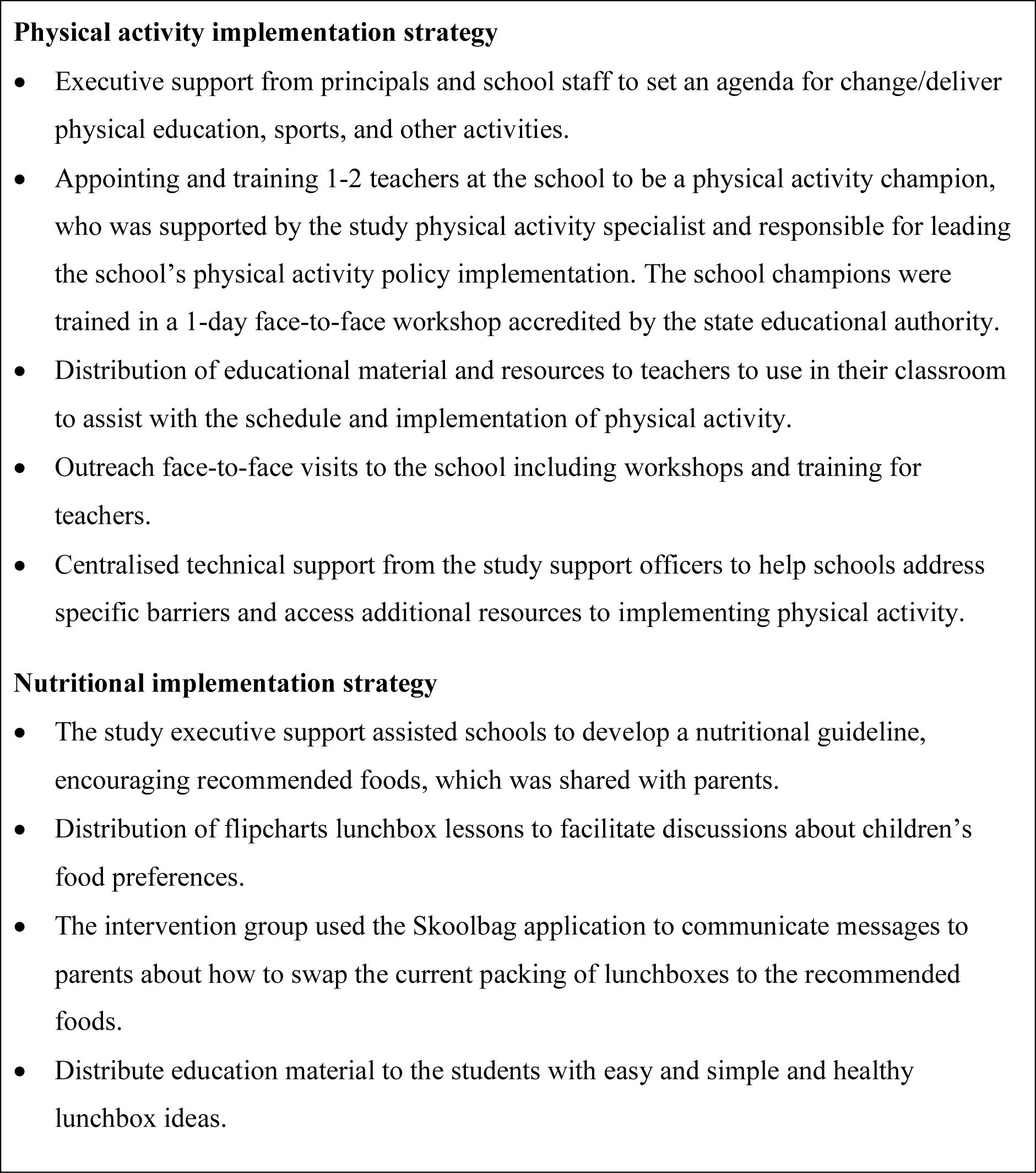
Brief description of the implementation strategies [1, 2].

### Outcomes

The primary outcome was an ordinal measure of pain frequency reported at 9-month follow-up as “often”, “sometimes”, or “never” over the last six months. The secondary outcomes were quality of life, pain severity, physical activity, sedentary time, prevalence of recent pain and pain-related impact. The outcomes were measured at baseline and post-intervention (nine months after baseline). **Table 1** describes details of primary and secondary outcomes.

**Table 1.** Primary and Secondary Outcomes.

| Outcomes | Outcomes details | Timepoint |
| --- | --- | --- |
| **Pain frequency** | The question “*Have you had pains or aches in your body?*” was adapted from the Young Spine Questionnaire.[32] We collapsed four response options into three by combining the response options ‘once in a while’ and ‘once or twice in total’ in a ‘sometimes’ category. Therefore, the response options analysed were ‘often’, ‘sometimes’, and ‘never’. | BL |
| **In the last 6 months** | “*In the past SIX months, have you had aches or pains in your body such as backache, headache or pain in your joints, bones or muscles. This does not include feelings such as overstretching or being hit by a ball during any sports.*” | FU |
| **Quality of Life** | PedsQL 4.0 child self-report (aged 8 to 12)[33-36] |  |
| **Total score** | 23-item instrument to measure health-related quality of life in paediatric populations including children physical, emotional, social, and school functioning during the past 1 month. Response options are in a 5-item Likert scale: “never”, “almost never”, “sometimes”, “often” and “almost always”. | BL, FU |
| **Functional score** | Reflects the physical function consequences of the chronic diseases in children. Higher scores indicate better health related quality of life. | BL, FU |
| **Pain severity** | Faces Pain Scale-Revised (FPS-R), a valid self-reported instrument to measure pain severity in children aged 4-16 years.[37-39] Continuous measure presented on a 6-point scale from 1 (no pain) to 6 (a lot of pain). “The faces below show how much something can hurt. The pain ranges from “No pain” to “A lot of pain”. Put a cross (X) on the face which shows how much pain you have had when it was worst.” | BL, FU |
| **Physical activity** | A continuous measure of minutes per day spent in moderate to vigorous-intensity physical activity (MVPA) during school days/hours only (i.e., Monday-Friday, 9am-3pm). The ranges of counts were derived from accelerometer (ActiGraph GT3X+, ActiGraph Corporation, Pensacola, FL) data as $\geq 262$ for the vertical axis wrist cut points.[40] | BL, FU |
| **Sedentary time** | Minutes per day spent sitting, lying down, and standing during school days/hours only (i.e., Monday-Friday, 9am-3pm). The ranges of counts were derived from accelerometer (ActiGraph GT3X+, ActiGraph Corporation, Pensacola, FL) data as 0-105 for the vertical axis wrist cut points.[40] | BL, FU |
| **Prevalence of recent pain** | A dichotomous measure asking students if they had aches or bodily pain “today” and “last week”. Adapted from the Young Spine Questionnaire.[32] |  |
| **Today** | For the analysis of this measure, we dichotomized the response options in yes/no by collapsing the ordinal response options “often”, “once in a while” and “once or twice in total” in the “yes” category.<br>*“Have you had any of these aches or pains in your body **today**? (Y/N)”* | BL, FU |
| **Last week** | *“Have you had any of these aches or pains in your body **in the last week**? (Y/N)”* | BL, FU |
| **Pain related impact** | Dichotomous measure asking students if bodily pain had stopped them from doing usual physical activities, going to school or any other usual activities, and if they had taken medication or sought health care because of their pain. The questions were adapted from a study in adolescents with low back pain.[41] For the analysis, we dichotomized the response options to yes/no by collapsing the ordinal response options “often”, “once in a while” and “once or twice in total” in the “yes” category. |  |
| **Physical activity** | *“Has your pain sometimes stopped you from doing sports or physical activities (e.g., running, cycling)?” (Y/N)* | BL, FU |
| **Usual activities** | *“Has your pain interfered with your normal activity? For example, carrying your school bag or doing up your shoes, walking upstairs.” (Y/N)* | BL, FU |
| **School attendance** | *“Have you stayed home from school because of the pain?” (Y/N)* | BL, FU |
| **Care seeking** | *“Have you been to a doctor, chiropractor, or physiotherapist because of the pain?” (Y/N)* | BL, FU |
| **Medication use** | *“Have you taken any medicine to relieve the pain?” (Y/N)* | BL, FU |

BL: baseline, FU: follow-up

### Statistical analysis

We conducted our analyses according to a preplanned statistical analysis plan published on Open Science Framework, following the intention to treat principle.[29] An independent statistician, blinded to group status and not involved with the study, conducted the analyses using SAS 9.4 (SAS Institute, Cary, North Carolina). The statistical analysis and presentation are consistent with the CHecklist for statistical Assessment of Medical Papers (CHAMP).[42]

### Power

The sample size was calculated to assess the main effect of physical activity intervention.[27] For this secondary analysis, a sample of 12 schools (6 per arm), with an average of 54 students per school, provided 80% power with an alpha level of 5% and an intraclass correlation of 0.03 to detect a difference where, at follow up, the control group is expected to have 25% pain never, 60% pain sometimes and 15% pain often and the treatment group 40% never, 52% sometimes and 8% often.

### Missing data

We investigated patterns of missing data for each outcome variable, which became a binary variable (missing/not missing). We used logistic mixed regression models, with random intercepts for school and class, to account for the two-stage clustering. We used multiple imputations (fully conditional specifications method) to handle missing data using Rubin’s rule.[43]

### Analyses

For the primary outcome, we used an ordinal mixed model univariable logistic regression to estimate the effect of the physical activity intervention on the pain frequency at follow-up, measuring the odds of reporting a lower frequency of pain. The proportional odds assumption was examined by running two logistic models for the two adjacent levels (student classrooms within schools), checking if the confidence intervals for the effect of the physical activity intervention overlapped, and running a non-mixed ordinal model, using the score test to test for unequal slopes. For the secondary outcomes, binomial mixed models were used to estimate the effect of the physical activity intervention on pain frequency in the last week and pain-related impact. Linear mixed models were used to estimate the effect of the physical activity intervention on daily activity, pain intensity, and quality of life. The physical activity intervention group was included as a fixed effect as well as the nutritional intervention group to account for possible effects of this second intervention strategy as part of the factorial design of the study. School ID was included as a random effect to account for the cluster design. Class ID was also included as a random effect when variation was detected between classes.[29]

We conducted three preplanned subgroup analyses for primary and secondary outcomes. These analyses assessed the differential effects of the treatment (physical activity support) for participants who, at baseline, had 1) pain in the last week, 2) primary pain location in the knee, and 3) pain location in the back. In addition to the fixed and random terms specified above, an additional fixed effect was included representing the subgroup of interest and an interaction term between the subgroup of interest and the treatment group.

We performed two sensitivity analyses. These included i) a pre-planned analysis of the primary outcome including participants with completed outcome data only, and ii) a post-hoc sensitivity analysis using a dichotomized primary outcome where response options “sometimes” and “never” were combined as infrequent pain and the response option “often” as frequent pain.[29]

## Results

From 2147 consenting students from the 12 schools in the original study, 815 (38%) students from grades 4, 5, and 6 returned a baseline survey. Seven hundred and nine children reported whether they had pain or not, and 633 indicated that they experienced pains or aches in their body at least “once or twice”. Of these children, 378 participants from schools randomised to the intervention group (physical activity) and 255 from schools randomised to the control group (nutrition support strategy only and waitlist). Baseline characteristics are presented in **Table 2**. At follow-up, 298 (78.9%) and 195 (76.5%) students from intervention and control groups provided pain frequency data (**Figure 2**).

**Figure 2:**
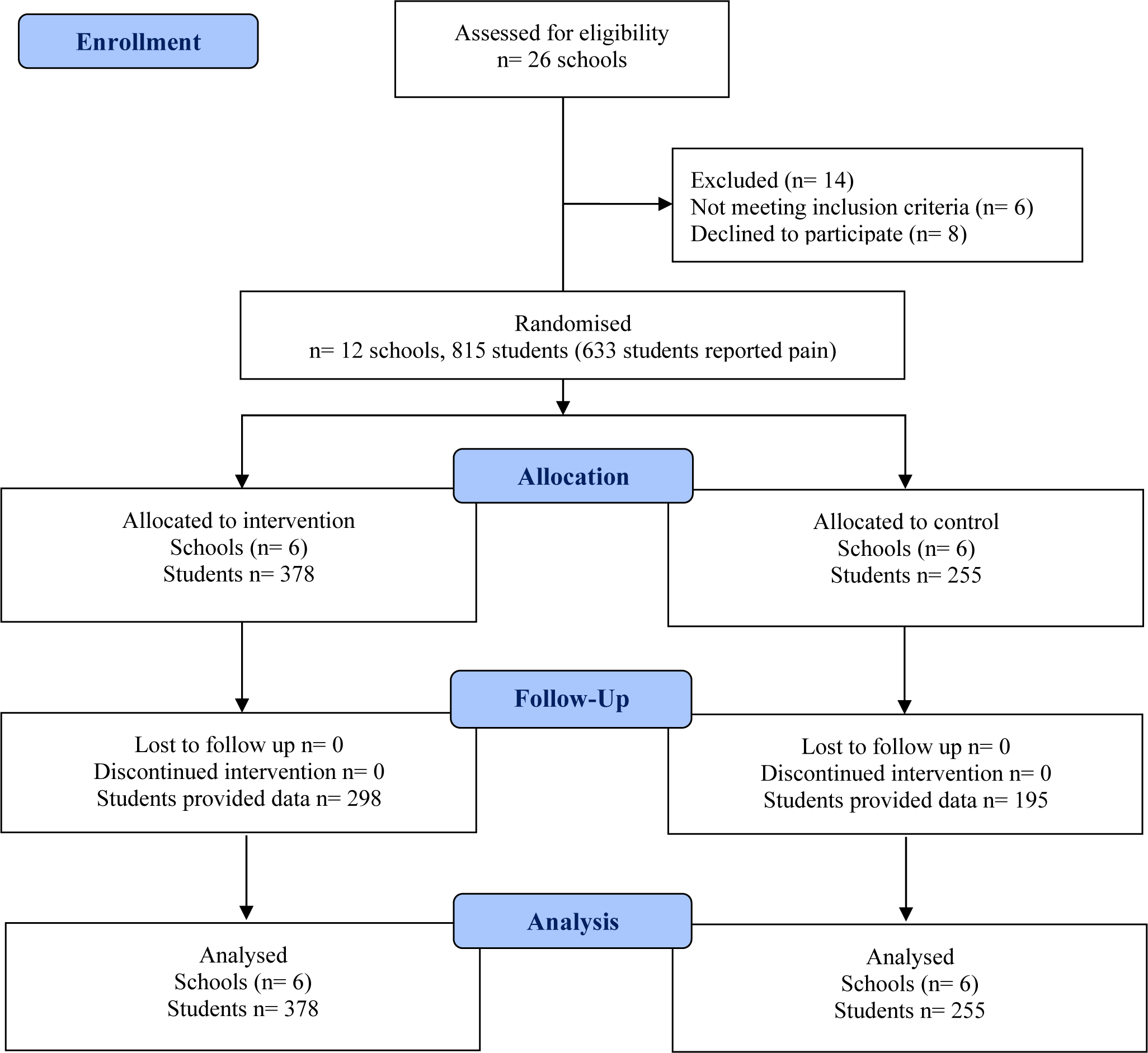
CONSORT flow chart of participants.

**Table 2.**
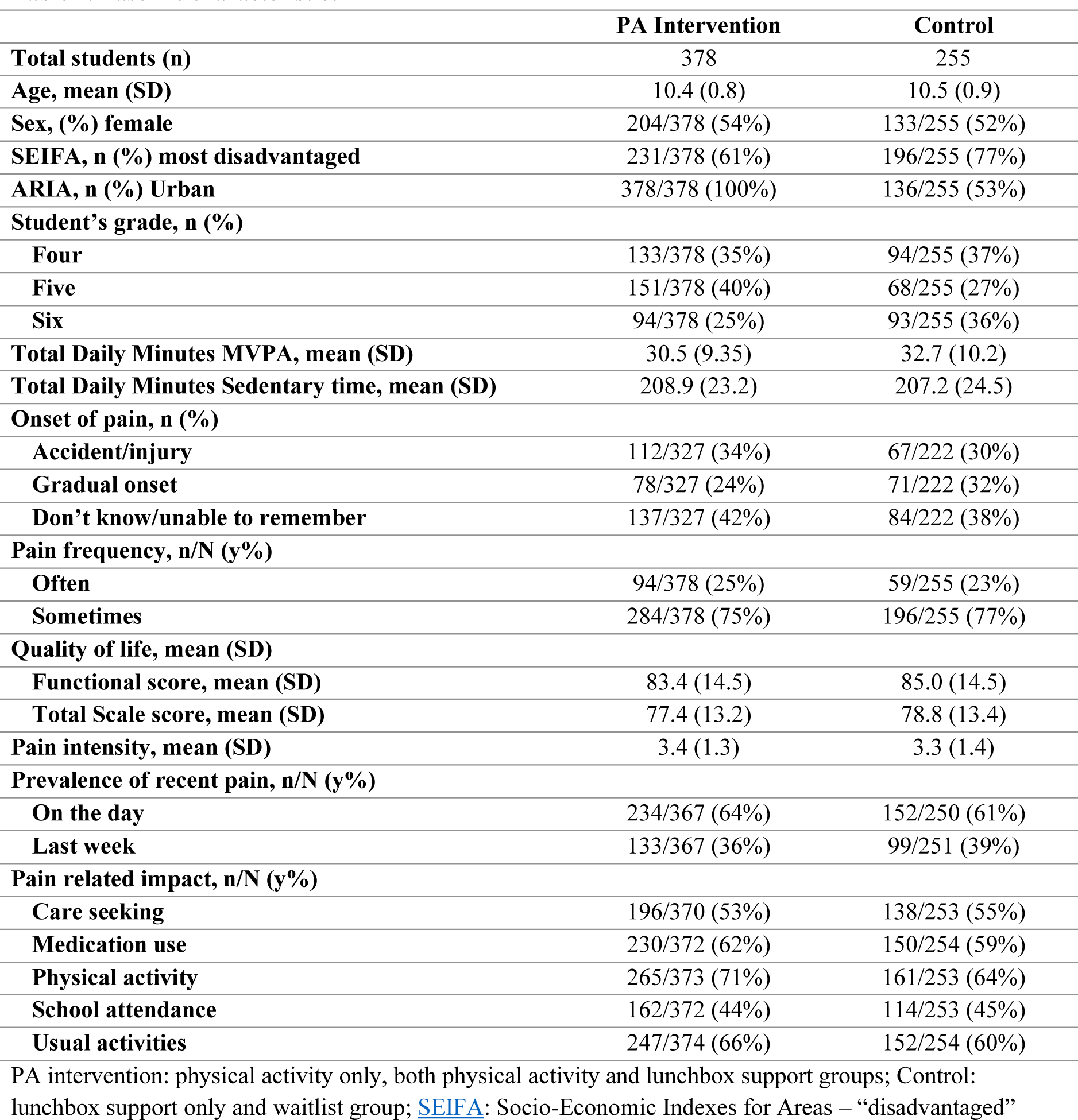
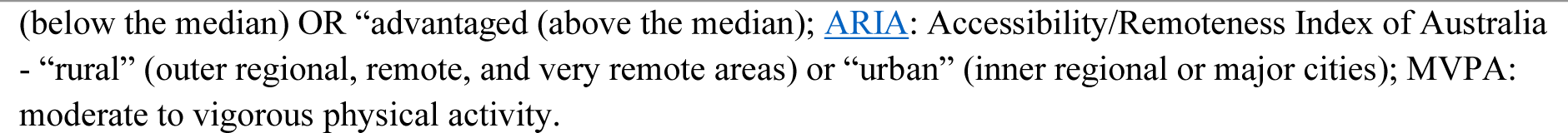
Baseline characteristics.

|  | **PA Intervention** | **Control** |
| --- | --- | --- |
| **Total students (n)** | 378 | 255 |
| **Age, mean (SD)** | 10.4 (0.8) | 10.5 (0.9) |
| **Sex, (%) female** | 204/378 (54%) | 133/255 (52%) |
| **SEIFA, n (%) most disadvantaged** | 231/378 (61%) | 196/255 (77%) |
| **ARIA, n (%) Urban** | 378/378 (100%) | 136/255 (53%) |
| **Student’s grade, n (%)** |  |  |
| **Four** | 133/378 (35%) | 94/255 (37%) |
| **Five** | 151/378 (40%) | 68/255 (27%) |
| **Six** | 94/378 (25%) | 93/255 (36%) |
| **Total Daily Minutes MVPA, mean (SD)** | 30.5 (9.35) | 32.7 (10.2) |
| **Total Daily Minutes Sedentary time, mean (SD)** | 208.9 (23.2) | 207.2 (24.5) |
| **Onset of pain, n (%)** |  |  |
| **Accident/injury** | 112/327 (34%) | 67/222 (30%) |
| **Gradual onset** | 78/327 (24%) | 71/222 (32%) |
| **Don’t know/unable to remember** | 137/327 (42%) | 84/222 (38%) |
| **Pain frequency, n/N (y%)** |  |  |
| **Often** | 94/378 (25%) | 59/255 (23%) |
| **Sometimes** | 284/378 (75%) | 196/255 (77%) |
| **Quality of life, mean (SD)** |  |  |
| **Functional score, mean (SD)** | 83.4 (14.5) | 85.0 (14.5) |
| **Total Scale score, mean (SD)** | 77.4 (13.2) | 78.8 (13.4) |
| **Pain intensity, mean (SD)** | 3.4 (1.3) | 3.3 (1.4) |
| **Prevalence of recent pain, n/N (y%)** |  |  |
| **On the day** | 234/367 (64%) | 152/250 (61%) |
| **Last week** | 133/367 (36%) | 99/251 (39%) |
| **Pain related impact, n/N (y%)** |  |  |
| **Care seeking** | 196/370 (53%) | 138/253 (55%) |
| **Medication use** | 230/372 (62%) | 150/254 (59%) |
| **Physical activity** | 265/373 (71%) | 161/253 (64%) |
| **School attendance** | 162/372 (44%) | 114/253 (45%) |
| **Usual activities** | 247/374 (66%) | 152/254 (60%) |

PA intervention: physical activity only, both physical activity and lunchbox support groups; Control: lunchbox support only and waitlist group; SEIFA: Socio-Economic Indexes for Areas – “disadvantaged”
(below the median) OR “advantaged (above the median); ARIA: Accessibility/Remoteness Index of Australia - “rural” (outer regional, remote, and very remote areas) or “urban” (inner regional or major cities); MVPA: moderate to vigorous physical activity.

### Primary and secondary outcomes

Estimates for primary and secondary outcomes are presented in **Table 3**. There was no difference in frequency of pain between groups (OR 1.0, 95% CI: 0.6 to 1.6) for each level of pain frequency (“often”, “sometimes”, and “never”) at follow-up. Slightly fewer students reported “sometimes” pain in the intervention (49%) compared to the control group (53%), and the intervention group had more students reporting “often” pain (36%) compared to the control (33%). Both groups had a mean pain intensity of three points in the FPS-R. The intervention group had a higher total daily physical activity (mean difference: 3.1 min/day, 95% CI: −0.02 to 6.2) and lower total sedentary time (mean difference: −5.5 min/day, 95% CI: −20.3 to 9.3). Between-group means were similar for the subscales of PedsQL 4 – total scale (0.5; 95% CI: −2 to 3) and physical functioning (−0.16; 95% CI: −2.9 to 2.5). We found lower pain-related impact on usual activities, but confidence intervals were wide (OR 0.6, 95% CI: 0.4 to 1.0).

**Table 3.** Primary and Secondary outcomes.

| Primary Outcome | PA Intervention | Control | Mean Difference (MD) or Odds Ratio (OR) (95% CI) | p-value |
| --- | --- | --- | --- | --- |
| **Pain frequency n/N (y%)** |  |  |  |  |
| Never | 45/298 (15%) | 27/195 (14%) | OR 1.0 (0.60, 1.64) | 0.987 |
| Sometimes | 147/298 (49%) | 104/195 (53%) |  |  |
| Often | 106/298 (36%) | 104/195 (33%) |  |  |
| **Secondary Outcomes** |  |  |  |  |
| **Quality of life** |  |  |  |  |
| Total score, mean (SD) | 78.0 (13.1) | 78.8 (13.1) | MD 0.52 (-2.0, 3.04) | 0.686 |
| Functional score, mean (SD) | 83.8 (13.4) | 84.8 (12.0) | MD -0.16 (-2.87, 2.55) | 0.907 |
| Pain severity, mean (SD) | 3.0 (1.4) | 3.0 (1.4) | MD -0.13 (-0.41, 0.16) | 0.389 |
| **Mean daily activity** |  |  |  |  |
| Total Daily Minutes MVPA, mean (SD) | 35.6 (11.7) | 32.6 (223, 10.3) | MD 3.08 (-0.02, 6.17) | 0.051 |
| Total Daily Minutes Sedentary time, mean (SD) | 194.0 (351, 25.2) | 201.7 (223, 30.2) | MD -5.52 (-20.31, 9.27) | 0.464 |
| **Prevalence of recent pain n/N (y%)** |  |  |  |  |
| On the day | 112/296 (38%) | 79/198 (40%) | OR 0.86 (0.53, 1.37) | 0.517 |
| Last week | 206/301 (68%) | 137/196 (70%) | OR 0.91 (0.60, 1.36) | 0.637 |
| **Pain related impact n/N (y%)** |  |  |  |  |
| Care seeking | 165/301 (55%) | 112/199 (56%) | OR 0.80 (0.42, 1.56) | 0.518 |
| Medication use | 184/299 (62%) | 122/199 (61%) | OR 1.07 (0.56, 2.04) | 0.833 |
| Physical activities | 204/299 (68%) | 134/198 (68%) | OR 0.90 (0.53, 1.52) | 0.692 |
| **School attendance** | 131/301 (44%) | 90/198 (45%) | OR 0.85 (0.35, 2.06) | 0.720 |
| **Usual activities** | 130/301 (43%) | 99/199 (50%) | OR 0.66 (0.42, 1.02) | 0.064 |

MVPA: moderate to vigorous physical activity

### Sensitivity and subgroup analyses

Sensitivity and subgroup analyses are presented in **Table 4**. The pre-planned analysis of the primary outcome, including only the complete cases, showed children in the control group had 1.23 times the odds of experiencing frequent pain, but CIs indicate a possible 32% reduction up to 220% increase in odds (95% CI 0.6 to 2.2). The post hoc analysis of pain frequency, dichotomised as infrequent and frequent, showed a small but uncertain increase in the odds of frequent pain in the intervention group (OR 1.09, 95% CI: 0.6 to 1.9).

**Table 4.**
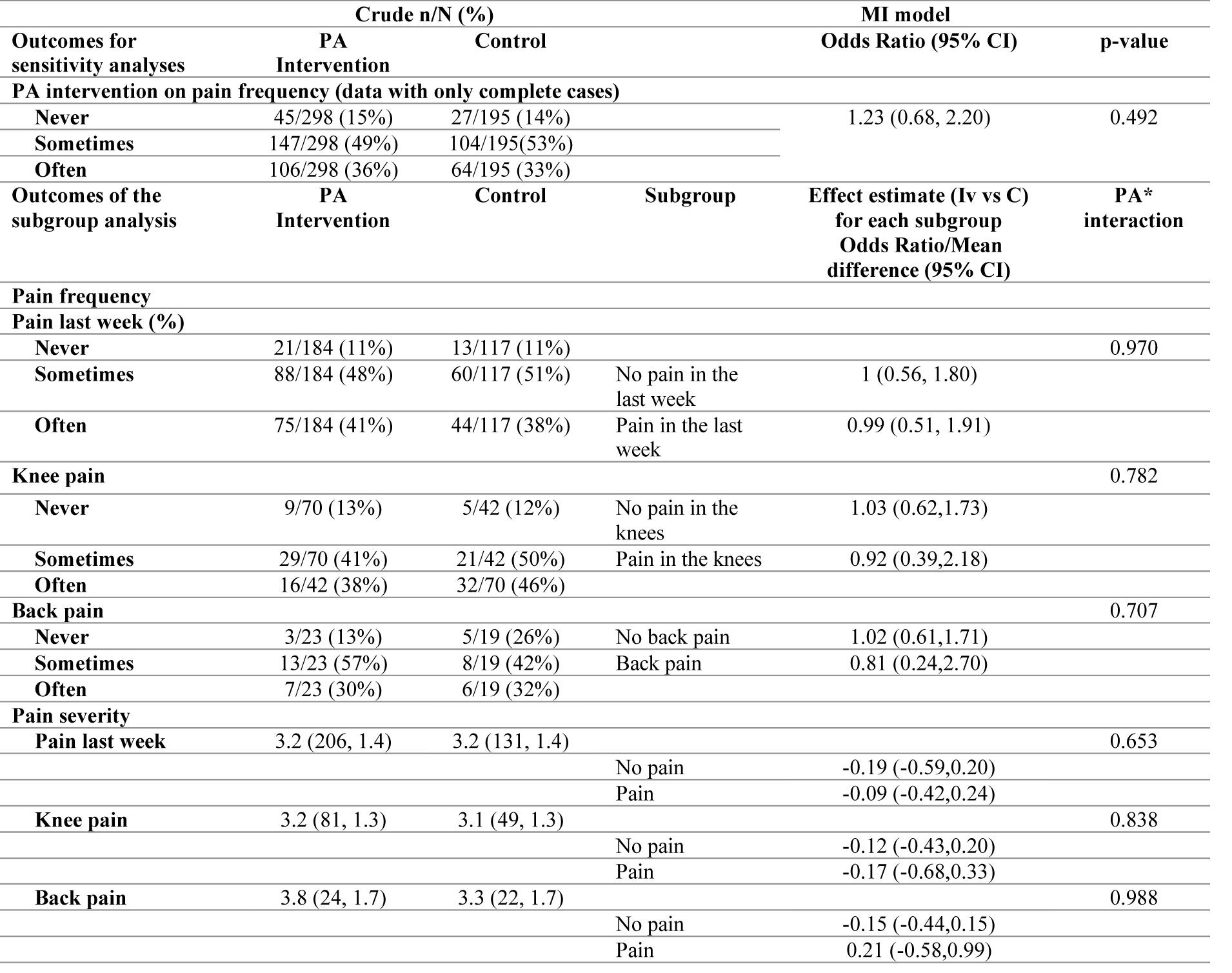
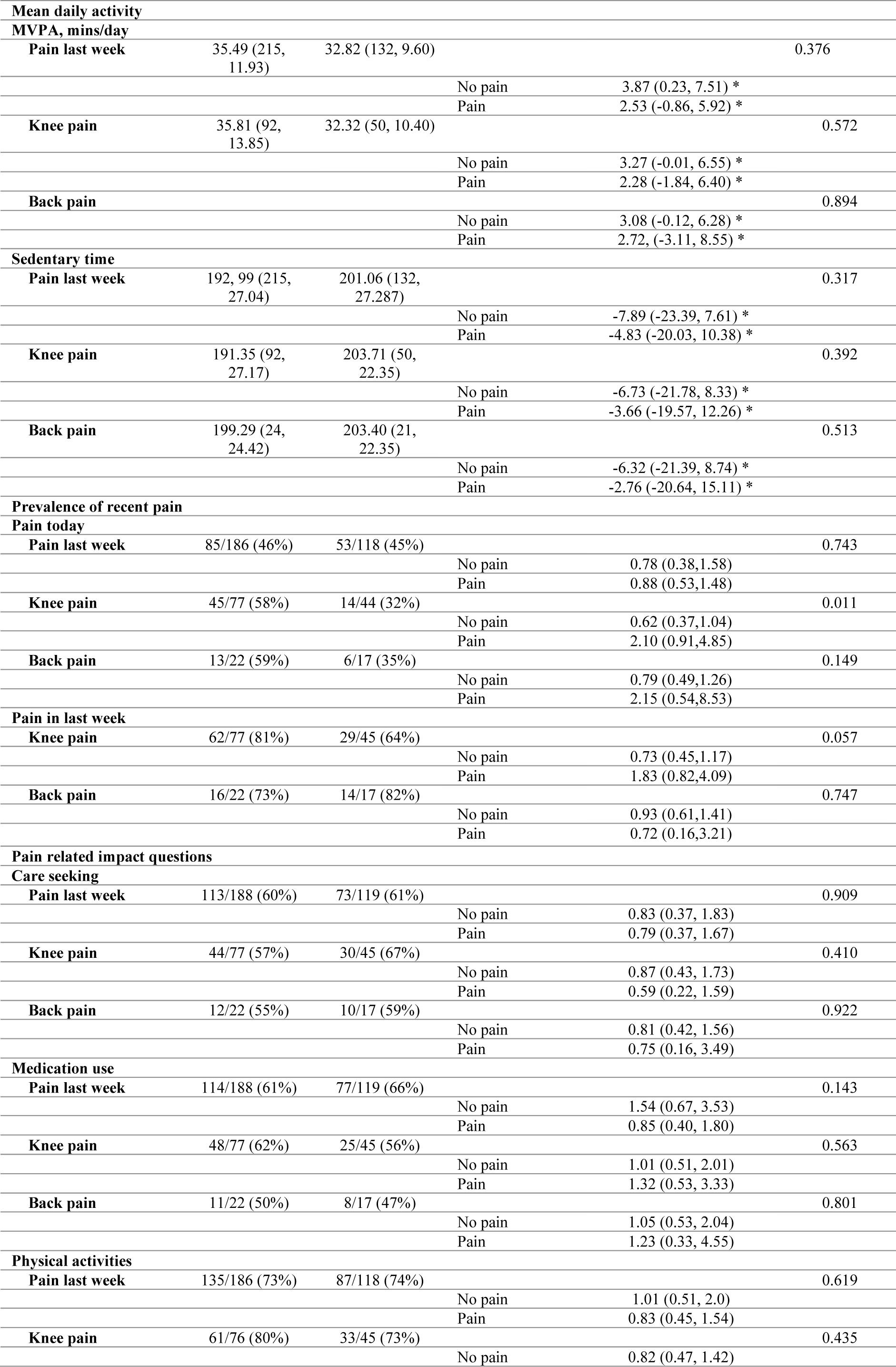
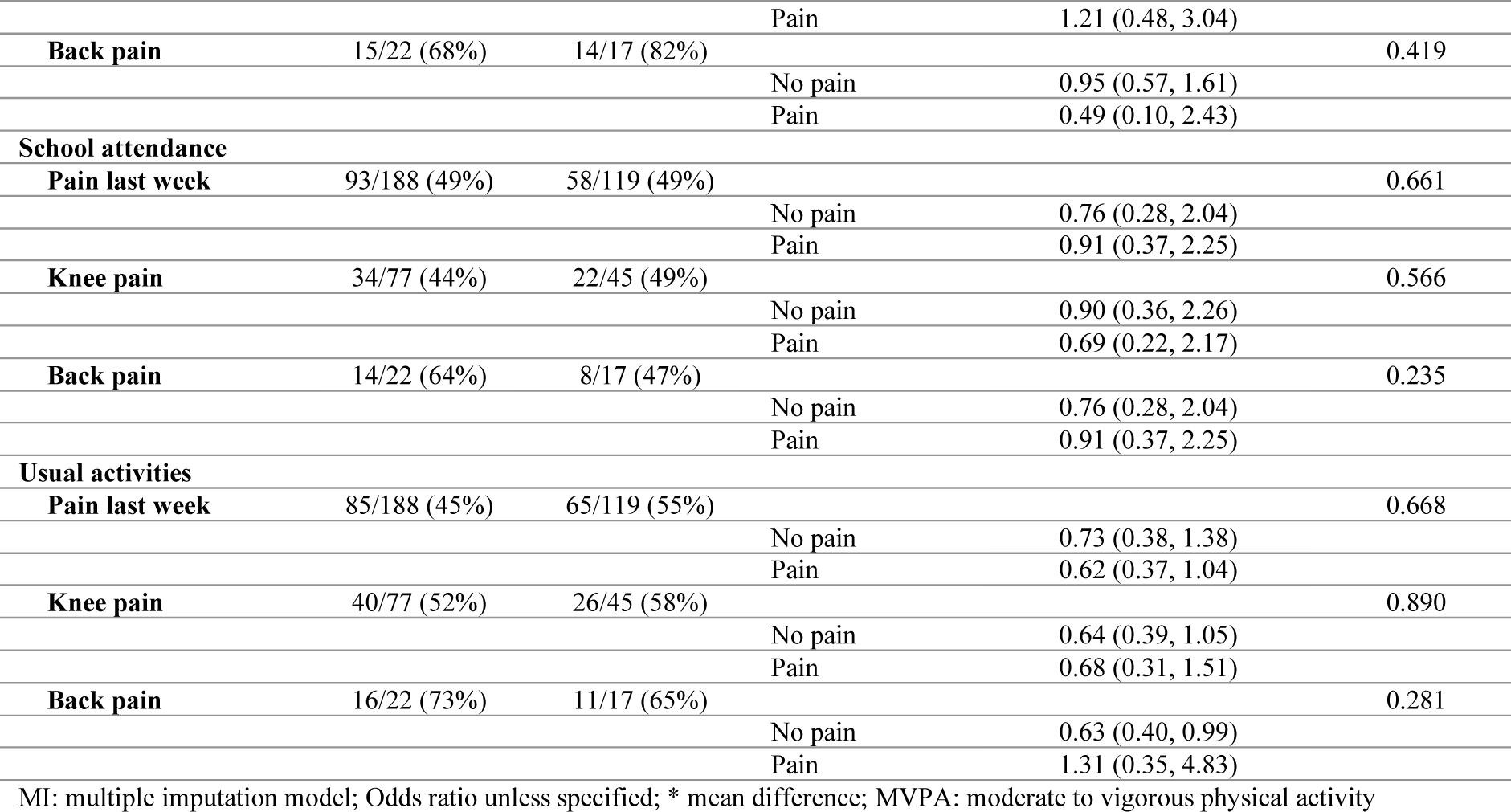
Sensitivity and Subgroup analyses.

The subgroup analysis showed that there was a slightly larger positive effect of the intervention on physical activity for students who had no pain in the last week (mean additional daily mins of MVPA 3.9, 95% CI: 0.2, 7.5) compared to students who had pain (mean additional daily mins of MVPA 2.5, 95% CI −0.8 to 5.9), with the CI suggesting uncertainty in the effect for students with recent pain.

## DISCUSSION

Although physical activity is recommended for children with pain, we found supporting more scheduling of physical activity in schools did not improve pain frequency or most secondary outcomes in students with a previous experience of MSK pain. We observed a small positive effect of the intervention in increasing daily minutes of MVPA for students with MSK pain, which was equivalent to the effect observed in the main trial. We found that recent pain (‘pain in the last week’) may reduce the size of the intervention’s effect on physical activity. Students from intervention schools also reported a lower pain impact on usual activities. However, as indicated by wide confidence intervals, the effect could be between a 58% reduction and a 2% increased impact.

### Strengths and limitations

This study has several strengths. First, this was a large high-quality cluster randomised trial. Second, we followed a pre-published statistical analysis plan to ensure transparency of data analyses. Third, we measured physical activity using accelerometers[44]. The weaknesses of this study are related to retention rates and measurement instruments. Although there was 100% retention of the schools (clusters), 21% of students from the intervention and 24% of students from the control did not provide data at follow-up. We acknowledge that the instrument used to measure pain frequency and impact might not have been sensitive enough. However, there are a few validated options to assess the consequence of pain in school-aged children.[45] We used questions that have been used in previous research on adolescents with low back pain. However, the questions have not been validated in elementary school-aged children.[41]

### Implications

Contrary to previous evidence,[28] our study indicates that a focus on increasing physical activity in schools may not improve pain outcomes of students with prior MSK pain. Our recent Cochrane review found physical activity produces a small improvement in pain and disability compared to usual care for children with juvenile idiopathic arthritis.[46] However, the certainty of evidence was very low from only three small RCTs.[46] The results of our current trial indicate uncertainty about whether a general approach to physical activity implemented in schools has benefits for pain outcomes in children.

It is important to note that our intervention supported teachers to increase physical activity scheduling during school time for all children and did not include any specific education about physical activity in children with MSK pain. The mean total scheduled PA time was 136 mins per week in intervention schools, an increase of approximately 37 min/week compared to the control schools.[27] We observed only a small increase in device-measured physical activity (∼3min/day). We could not determine if this dose was insufficient or if focusing on physical activity alone is of no benefit to pain outcomes in school children.

We found that scheduling physical activity in the school setting might help children with MSK pain to be physically active despite MSK pain. We also found a potential reduction of the impact of pain on usual activities. Previous studies investigating the relationship between pain outcomes and physical activity demonstrated that MSK pain can predispose to sedentary time.[12, 44] Similarly to previous research,[47] we observed that students with recent pain are less active. Overall, these results suggest that interventions promoting physical activity can be beneficial to keep children with MSK pain active and reduce the impact of pain on usual activity, but children with recent pain may need additional support.

Future research could explore if providing teachers and students with more information about pain and physical activity leads to greater benefits. Other theories of human functioning and school organisation reveal that students rely on authority figures, such as parents and teachers, to support the adoption of healthier behaviours.[48] In addition, a child’s concept of pain is transferable from parents and teachers.[49, 50] Therefore, engaging parents and teachers to support children with MSK pain to become and stay physically active might result in better pain outcomes.

## CONCLUSIONS

Supporting schools to increase scheduling of physical activity did not improve pain frequency or other pain outcomes. However, the intervention resulted in a small increase in physical activity levels and reduced impact on usual activities in children with MSK pain. Beyond encouraging children to be active, the benefit of school-based physical activity for students with MSK pain is uncertain.

## Data Availability

All data produced in the present study are available upon reasonable request to the authors

